# Magnetic resonance spectroscopy of anoxic brain injury after cardiac arrest

**DOI:** 10.1101/2021.05.13.21257029

**Authors:** Jong Woo Lee, Lasya Sreepada, Matthew Bevers, Karen Li, Benjamin Scirica, Danuzia Santana da Silva, Galen V. Henderson, Camden Bay, Alexander P Lin

**Affiliations:** Department of Neurology, Brigham and Women’s Hospital, Boston, MA; Department of Radiology, Brigham and Women’s Hospital, Boston, MA; Department of Medicine, Division of Cardiology, Brigham and Women’s Hospital, Boston, MA

**Author notes:** **Corresponding Author**: Alexander P Lin. **Contributions**: Jong Woo Lee: Drafting/revision of the manuscript for content, including medical writing for content; Major role in the acquisition of data; Study concept or design; Analysis or interpretation of data Lasya Sreepada: Drafting/revision of the manuscript for content, including medical writing for content; Analysis or interpretation of data Matthew Bevers: Drafting/revision of the manuscript for content, including medical writing for content; Analysis or interpretation of data Karen Li: Analysis or interpretation of data Benjamin Scirica: Drafting/revision of the manuscript for content, including medical writing for content; Major role in the acquisition of data; Study concept or design Danuzia Santana da Silva: Drafting/revision of the manuscript for content, including medical writing for content; Major role in the acquisition of data Galen V. Henderson: Drafting/revision of the manuscript for content, including medical writing for content; Major role in the acquisition of data; Study concept or design Camden Bay: Drafting/revision of the manuscript for content, including medical writing for content; Analysis or interpretation of data Alexander P Lin: Drafting/revision of the manuscript for content, including medical writing for content; Major role in the acquisition of data; Study concept or design; Analysis or interpretation of data. **Supplemental**: Response to Reviewers Updated STROBE Checklist. **Statistical Analysis performed by**: Camden Bay, PhD.

**Keywords:** [290] Cardiac, [284] EEG, see Epilepsy/Seizures (S), [62] EEG, [120] MRI, [125] MRS

## Abstract

**Objective:** We describe magnetic resonance spectroscopy (MRS) changes in comatose patients undergoing targeted temperature management (TTM) after cardiac arrest, and their relationships to relevant clinical, MRI, and EEG variables.

**Methods:** A prospective cohort of 50 patients was studied. The primary outcome was coma recovery to follow commands. Comparison of MRS measures in the posterior cingulate gyrus, parietal white matter, basal ganglia, and brainstem were also made to 25 normative control subjects.

**Results:** Fourteen of 50 achieved coma recovery. Compared to patients who recovered, there was a significant decrease in total N-acetyl-aspartate (NAA/Cr) and glutamate; and an increase in lactate (Lac/Cr) and glutamine in patients who did not recover, with changes most prominent in the posterior cingulate gyrus. Patients who recovered had a decrease in NAA/Cr as compared to control subjects. Coma recovery was seen in patients with a moderate decrease in NAA/Cr, but the appearance of lactate resulted in a poor outcome. NAA/Cr had a linear relationship with MRI cortical apparent diffusion coefficient (ADC); lactate level exponentially increased with decreasing ADC. EEG suppression/burst suppression was universally associated with lactate elevation.

**Conclusions:** NAA and lactate changes are associated with clinical/MRI/EEG changes consistent with anoxic brain injury and are most prominent in the posterior cingulate gyrus. NAA/Cr decrease observed in patients with good outcomes suggests mild anoxic injury in patients asymptomatic at hospital discharge. The appearance of cortical lactate represents a deterioration of aerobic energy metabolism and is associated with EEG background suppression, synaptic transmission failure, and severe, potentially irreversible anoxic injury.

## Introduction

Anoxic brain injury represents the leading cause of mortality and long-term disability after cardiac arrest. More than 350,000 adults suffer out-of-hospital cardiac arrest annually in the United States, after which up to 80% remain comatose immediately thereafter.^1^ As clinical examination may not be sufficiently informative after targeted temperature management (TTM)^2^, ancillary tests have been utilized to assess prognosis, including EEG, somatosensory evoked potential (SSEP), MRI, and neuron specific enolase (NSE).

MR spectroscopy (MRS) has been utilized along with these other measures in assessing prognosis.^3^ Few studies have examined the range of changes observed in brain-specific metabolic markers through MRS in adult patients undergoing TTM. In neonatal patients undergoing TTM for hypoxic-ischemic injury due to birth asphyxia, changes in NAA and choline (Cho) levels, as measured by MRS, were associated with clinical outcome after TTM, though there is disagreement regarding the utility of the measurement of lactate (Lac) or the optimal brain region to be examined.^4, 5^ Furthermore, previous studies have focused on brain regions such as the pons, basal ganglia, thalamus, and corpus callosum which for technical reasons may not provide the most sensitive measure for MRS changes after cardiac arrest. The wide array of metabolites assessed by MRS allows for simultaneous assessment of both neuronal integrity and aerobic metabolism, both of which are primary pathologies in anoxic brain injury.

We examine changes of several metabolites, as measured by MRS, in patients comatose after cardiac arrest, comparing 1) patients with good vs poor neurological outcome; 2) patients with anoxic brain injury to normative control subjects in the posterior cingulate gyrus (PCG), parietal white matter (PWM), basal ganglia (BG), and brainstem (BS). These results are placed in the context of their EEG and conventional MRI changes to identify and quantify markers of anoxic brain injury after TTM and to determine brain regions that are preferentially affected in anoxic brain injury.

## Methods

### Patient cohort and clinical outcome

This is a single-center prospective study of patients who underwent TTM for coma after cardiac arrest and return of spontaneous circulation (ROSC) at the Brigham and Women’s Hospital between October 2016 and November 2020. Normative control subjects consisted of 25 athletic male volunteers without a history of head trauma or neurological disease, between ages 45 and 70 (mean 57.9 ± 7.0).

The following data points were collected: age, sex, and pulseless rhythm at the time of arrest (ventricular fibrillation/shockable vs pulseless electrical activity/asystole). Clinical exam findings included pupillary light response, corneal reflex, and motor response. Motor findings were dichotomized as flexor response or better (including flexion, withdrawal localization, or normal function) versus extensor response, triple-flexion, or no response. Motor function was scored based on the best response in either the upper or lower extremities.

The main clinical outcome was defined as ***coma recovery*** in those patients who regained the ability to open eyes and follow commands before hospital discharge; any response less than this was considered a poor outcome. As some patients were able to follow commands but were still cognitively impaired, we identified patients with a cognitive return to baseline. Cerebral performance category (CPC) at discharge were binarized to good (CPC 1-2) or poor (CPC 3-5) outcome.^6^

### Cardiac arrest protocol

TTM was performed according to the previously published local protocol.^7^ Eligible resuscitated cardiac arrest patients received TTM at 33°C or 36°C for 24 hours, using ice packs and a surface cooling device (Arctic Sun System, Medivance, Louisville, CO). Patients were intubated and sedated for 24 h with Propofol (0-83 mcg/kg/min) or midazolam (0-3 mg/h), and fentanyl (25 mcg/h). Cisatracurium (0.15 mg/kg IV q10min PRN, escalated to infusion at 0.5 mcg/kg/min for persistent shivering) was administered for shivering. After 24 hours, patients were rewarmed by 0.25°C/hour. When clinically feasible, sedation was lightened after return to normothermia for clinical assessments. Withdrawal of life-sustaining treatment (WLST) was performed on a case-by-case basis through informed decision-making by the patient’s family or guardian in close collaboration with the medical team and the patient’s family members.

### EEG

Continuous video EEG (cEEG) monitoring (Natus XLTEK system, Pleasanton, CA) was placed according to the international 10-20 system as soon as possible after starting TTM. Patients were recorded for a minimum of 24h after normothermia was achieved. EEG data were interpreted using the American Clinical Neurophysiology Society (ACNS) critical care EEG terminology^8^ and classified into 3 categories (highly malignant, malignant, benign) per Westhall et al.^9^ EEG assessment was made on the day of the MRI/MRS study.

### Neuroimaging

#### MRI

MRI and MRS scans were performed on a 3T Siemens Verio MRI scanner using a 32-channel head coil, on a mean of 6.4 ± 6.3 days (median 4.8 days) after cardiac arrest. Structural imaging included 3D T1-weighted (MPRAGE: 1×1×1 mm3, TR= 2530.0 ms, TE= 3.36 ms) and diffusion-weighted images. Regional analysis of ADC images was carried out using FSL (Oxford Center for Functional MRI of the Brain, Oxford, UK).^10^ ADC images were first co-registered to the T1 weighted image for each subject. The T1 images were then non-linearly registered to the Montreal Neurologic Institute (MNI) Brain Atlas, and the same transformation was applied to the ADC images. The success of automated registration was then manually confirmed. Artifacts were removed by filtering out signal intensity less than 200 mm^2^/s, while CSF was removed by filtering signal intensity greater than 2000 mm^2^/s. Regional anatomical masks were created using a 50% probability map from the MNI atlas. Mean ADC signal intensity was then calculated for each region. The percentage of whole brain and cortical voxels with ADC values < 650 mm^2^/s were similarly calculated from whole brain and cortical anatomical maps.

#### MRS

Due to limitations imposed by the local institutional review board, MRS was obtained only with a concurrent clinically indicated MRI. MRS was performed in four brain regions using a single voxel PRESS sequence (TE=30ms, TR=2000ms, 20×20×20 mm^3^, 128 averages). The use of a short echo time allows for the characterization of additional metabolites other than NAA, Cr, and Cho, such as glutamate, glutamine, and myoinositol. Previous studies utilized long echo times (TE > 144 ms) whereby these additional metabolites can no longer be detected due to their relatively fast relaxation times. Brain regions include the PCG, PWM, left BG, and BS as shown in Figure 1. The reason for selecting the PCG and PWM is that these regions have been shown to be highly sensitive to anoxic/hypoxic injury^11^ and are sites of reduced cerebral perfusion.^12^ The BG and BS were acquired so that they could be compared to other MRS studies in anoxia^3-5^. Anatomical landmarks from the T1-weighted images determined voxel locations, and subsequently, automated optimization (3D shimming, transmit gain, frequency adjustment, and water suppression) was performed on the voxel. When necessary, MRI technicians manually shimmed to a line width of <15 Hz of the full-width half maximum (FWHM) of the unsuppressed water spectrum in the basal ganglia and brainstem. Manual shimming was not required for the PCG or PWM as both of these regions have excellent B0 homogeneity. This is another advantage of utilizing these voxel locations. MRS of the BG (2 patients) and BS (12 patients) were not obtained due to time constraints.

**Figure 1:**
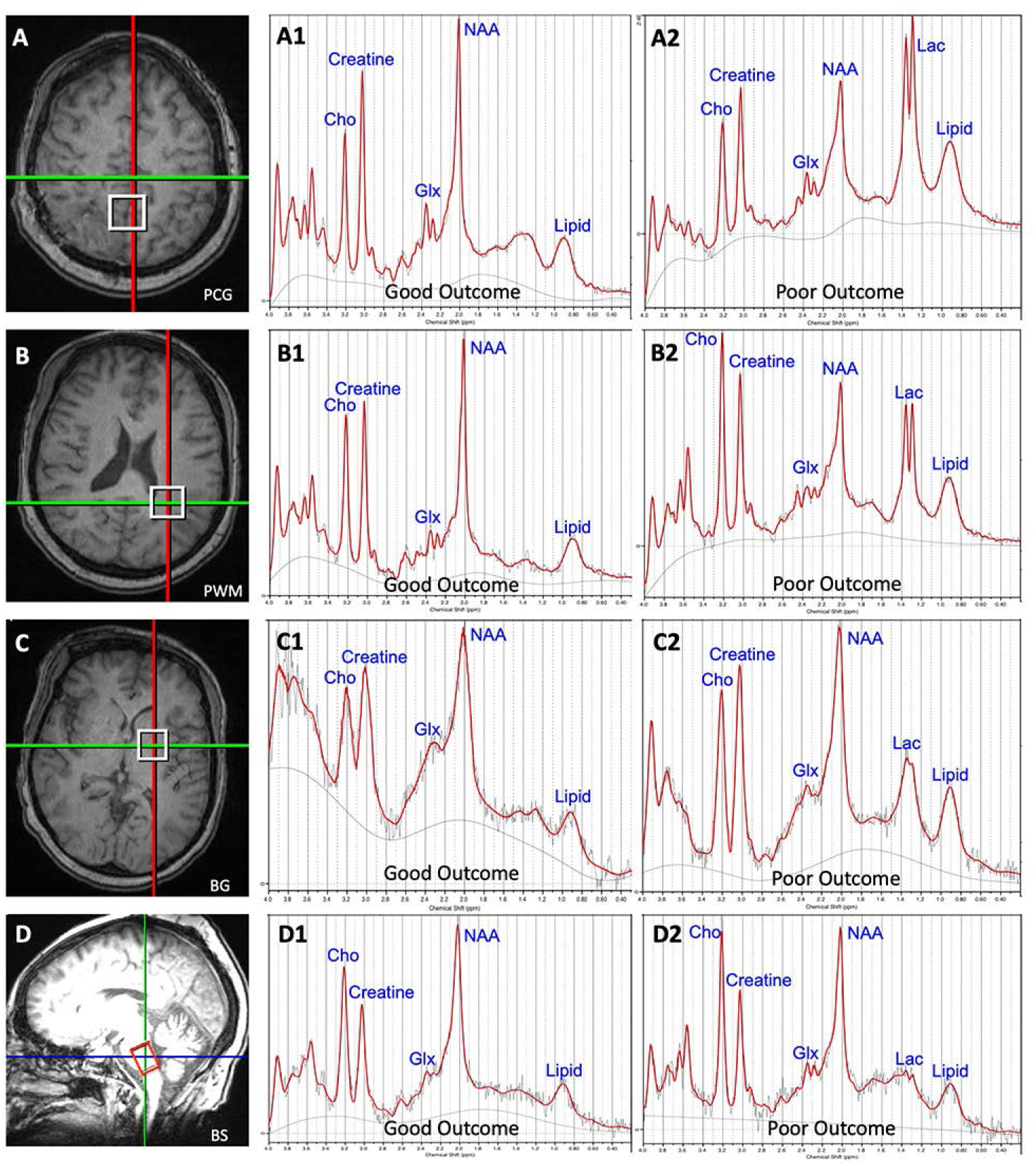
MRS voxel placement and representative spectra in patients with good and poor outcome Placement of the MRS voxels: A) Posterior cingulate gyrus (PCG); B) Posterior white matter (PWM); C) Basal ganglia (BG); D) Brainstem (BS). Spectra of illustrative patients with good outcomes (A1-D1) or poor outcomes (A2-D2).

Single-voxel MRS raw data were pre-processed using singular value decomposition (SVD)-based channel combination, spectral registration to correct for frequency drift, and water suppression using the Hankel SVD method and OpenMRSLab.^13^ Metabolites were then fit and quantified using linear combination models (LCModel)^14^ yielding the following measurements: total NAA (including NAA + NAA-Glutamate), total creatine (Cr, including Cr + PhosphoCr), total Cho (including PhosphoCho + Cho), Glutamate (Glu), Glutamine (Gln), Glutamate+Glutamine (Glx), myoinositol (mI), and lactate (Lac). Cr concentrations were not significantly different between the different cohorts and therefore used to normalize the data across the study. Normative control scans used for comparison were acquired using the same scan protocol (PCG and PWM) and on the same scanner.

In all subjects, major metabolites of NAA, Cr, Cho, and mI had Cramer-Rao Lower Bounds (CRLB) of less than 20%. Due to the high CRLB of Lac at very low concentrations, the data was not filtered to include subjects with low Lac to avoid any study bias.^15^

### Statistical Analysis

Comparative statistics (Student’s independent samples t-test, Fisher’s exact test, Wilcoxon rank-sum test) were used as appropriate. Statistically significant results were evaluated using a multivariable logistic regression model with dichotomized coma recovery as well as CPC at hospital discharge (1-2 vs 3-5) as dependent variables. Correlations between variables of interest were assessed using Spearman’s correlation with 95% confidence intervals. Multiple comparisons were adjusted for using a false discovery rate set at 0.05.^16^ P-values presented in the manuscript text are the raw p-values; all p-values are also presented in Tables 1 and 2 with an indication of their statistical significance after adjustment for multiple comparisons. Receiver operating characteristic (ROC) analyses were performed to obtain discrimination thresholds. Calculations were performed using R 3.3.2 (www.R-project.org).

**Table 1:** Characteristics of the study population.

|  | No coma recovery<br>(n=36) | Coma recovery<br>(n=14) | p-value |
| --- | --- | --- | --- |
| **Age in years (standard deviation)** | 61.2 (11.9) | 60.1 (15.0) | 0.81 |
| **Sex** |  |  | 0.52 |
| M | 21 (58.3%) | 10 (71.4%) |  |
| **†Pulseless rhythm** |  |  | 0.31 |
| VT/VF | 9 (25.7%) | 6 (42.9%) |  |
| Non-shockable | 26 (74.3%) | 8 (57.1%) |  |
| **CPC at discharge** |  |  | <0.001* |
| 1-2 | 0 | 12<br>CPC1=4<br>CPC2=8 |  |
| 3-5 | 36<br>CPC4=3<br>CPC5=33 | 2<br>CPC3=1<br>CPC5=1 |  |
| **‡CPC at 3-6 months** |  |  | <0.001* |
| 1-2 | 0 | 10<br>CPC1=6<br>CPC2=4 |  |
| 3-5 | 36<br>CPC4=1<br>CPC5=35 | 3<br>CPC5=3 |  |
| Days to WLST (interquartile range) | 6 (6) |  |  |
| **§Target temperature** |  |  | 0.52 |
| 33 | 31 (88.9%) | 11 (84.7%) |  |
| 36 | 4 (11.1%) | 2 (15.4%) |  |
| **Clinical exam** |  |  |  |
| Pupillary reflexes (%) | 26 (72.2%) | 14 (100%) | 0.045 |
| Corneal reflexes (%) | 20 (55.6%) | 14 (100%) | 0.002* |
| Motor exam flexion or better (%) | 4 (11.1%) | 14 (100%) | <0.001* |
| **MRI** |  |  |  |
| Median % whole brain voxels with ADC below 650 mm<sup>2</sup>/s (range) | 22.9 (1.2-95.9) | 2.5 (0.9-6.1) | <0.001* |
| Median % cortical voxels with ADC under 650 mm<sup>2</sup>/s (range) | 19.6 (1.2-95.6) | 2.1 (0.6-4.6) | <0.001* |
| Whole brain ADC (mean, sd) | 869.7 (157.7) | 998.7 (60.5) | <0.001* |
| Cortex ADC (mean, sd) | 923.3 (108.7) | 1080.4 (78.1) | <0.001* |
| White matter ADC (mean, sd) | 777.1 (142.8) | 828.3 (49.8) | <0.001* |
| Acc ADC (mean, sd) | 835.6 (156.7) | 880.7 (102.1) | 0.24 |
| Caudate ADC (mean, sd) | 901.0 (170.4) | 969.2 (116.0) | 0.12 |
| Putamen ADC (mean, sd) | 717.6 (123.9) | 796.4 (46.6) | 0.002* |
| Thalamus ADC (mean, sd) | 880.0 (128.8) | 925.0 (46.7) | 0.076 |
| Basal ganglia ADC (mean, sd) | 842.7 (130.7) | 905.0 (73.2) | 0.039 |
| Amygdala ADC (mean, sd) | 980.6 (158.8) | 1002.1 (77.9) | 0.53 |
| Hippocampus ADC (mean, sd) | 995.7 (175.2) | 1089.0 (116.3) | 0.035 |
| Cerebellum ADC (mean, sd) | 838.9 (150.5) | 929.4 (82.7) | 0.010* |
| **EEG on day of MRI/MRS** |  |  |  |
| Continuous/nearly continuous | 7 (19.4%) | 14 (100%) | <0.001* |
| Discontinuous, burst suppressed,<br>or completely suppressed | 29 (80.6%) | 0 (0%) |  |
| Generalized periodic discharges | 16 (44.4%) | 9 (64.3%) | 0.35 |
| Myoclonus | 6 (16.7%) | 4 (28.6%) | 0.44 |
| Status epilepticus | 6 (16.7%) | 2 (14.3%) | 1 |
| \|\|Reactivity-yes | 8 (33.3%) | 10 (100%) | <0.001* |
| Reactivity-no | 16 (66.7%) | 0 (0%) |  |
| Somatosensory evoked potential |  |  |  |
| N20 present at least one limb | 2 | 1 |  |
| Absent | 1 | 0 |  |
| Neuron specific enolase |  |  |  |
| >33ng/mL | 6 | 1 |  |
| <33ng/mL | 2 | 2 |  |

\*Statistically significant after correction for multiple comparisons (FDR=0.05). †Rhythm
unknown in one patient; ‡One patient lost to follow-up. §Fever avoided in 2 patients but did not
undergo TTM; ||Reactivity not tested or ascertainable in 16 patients. P-values not calculated for
somatosensory evoked potential studies and neuron specific enolase due to the small number
of subjects

**Table 2:** MRS peaks vs outcome.

|  | No coma<br>recovery (n=30)<br>Median (IQR) | Coma recovery<br>(n=12)<br>Median (IQR) | p-value (Wilcoxon) |
| --- | --- | --- | --- |
| **PCG** |  |  |  |
| NAA+NAAG/Cr | 0.82 (0.29) | 1.26 (0.17) | <0.001* |
| Lac/Cr | 0.39 (0.51) | 0.04 (0.06) | <0.001* |
| Glu/Cr | 0.90 (0.25) | 1.12 (0.24) | 0.007* |
| Gln/Cr | 1.01 (0.75) | 0.49 (0.23) | <0.001* |
| Glx/Cr | 2.10 (0.63) | 1.56 (0.40) | 0.002 |
| MI/Cr | 0.55 (0.23) | 0.69 (0.15) | 0.028 |
| **PWM** |  |  |  |
| NAA+NAAG/Cr | 1.48 (0.33) | 1.71 (0.19) | 0.016 |
| Lac/Cr | 0.22 (0.23) | 0.08 (0.11) | 0.005 |
| Glu/Cr | 0.83 (0.17) | 0.83 (0.23) | 0.91 |
| Gln/Cr | 0.47 (0.37) | 0.41 (0.16) | 0.19 |
| Glx/Cr | 1.36 (0.41) | 1.30 (0.45) | 0.24 |
| MI/Cr | 0.80 (0.25) | 0.77 (0.11) | 0.89 |
| **BG** |  |  |  |
| NAA+NAAG/Cr | 0.75 (0.19) | 0.73 (0.15) | 0.92 |
| Lac/Cr | 0.14 (0.18) | 0.015 (0.080) | 0.01 |
| Glu/Cr | 0.90 (0.19) | 0.95 (0.16) | 0.94 |
| Gln/Cr | 0.85 (0.57) | 0.65 (0.22) | 0.06 |
| Glx/Cr | 1.78 (0.61) | 1.53 (0.41) | 0.040 |
| MI/Cr | 0.53 (0.24) | 0.62 (0.34) | 0.37 |
| **BS** |  |  |  |
| NAA+NAAG/Cr | 1.49 (0.33) | 1.71 (0.19) | 0.019 |
| Lac/Cr | 0.22 (0.23) | 0.078 (0.11) | 0.007 |
| Glu/Cr | 0.83 (0.17) | 0.83 (0.23) | 0.98 |
| Gln/Cr | 0.48 (0.36) | 0.41 (0.16) | 0.17 |
| Glx/Cr | 1.40 (0.42) | 1.30 (0.45) | 0.20 |
| MI/Cr | 0.79 (0.25) | 0.77 (0.11) | 0.97 |

\*Statistically significant after correction for multiple comparisons (FDR=0.05); Glu: glutamine;
Glu: glutamate; Glx: Glu+Gln

**Table 3:** ROC and cut-off values.

|  | **AUC** | **Optimal cut-off** | **Sensitivity and specificity at optimal cut-off** | **Cut-off required for 100% sensitivity for coma recovery; specificity** |
| --- | --- | --- | --- | --- |
| **PCG** |  |  |  |  |
| NAA/Cr | 0.96 | 1.11 | 85.7%, 88.9% | 0.98, 77.8% |
| Lac/Cr | 0.91 | 0.14 | 83.3%, 92.9% | 0, 0% |
| Lac/NAA | 0.93 | 0.12 | 86.1%, 100.0% | 0, 0% |
| **PWM** |  |  |  |  |
| NAA/Cr | 0.72 | 1.52 | 85.7%, 61.1% | 1.23, 19.4% |
| Lac/Cr | 0.76 | 0.16 | 66.7, 85.7% | 0, 0% |
| Lac/NAA | 0.77 | 0.091 | 65.7%, 85.7% | 0, 0% |
| **BG** |  |  |  |  |
| NAA/Cr | 0.48 | 0.73 | 55.6%, 47.1% | 0.64, 23.5% |
| Lac/Cr | 0.81 | 0.112 | 70.6%, 88.9% | 0, 0% |
| Lac/NAA | 0.78 | 0.11 | 76.5%, 77.8% | 0, 0% |
| **BS** |  |  |  |  |
| NAA/Cr | 0.56 | 1.92 | 50.0%, 76.0% | 0.99, 8.0% |
| Lac/Cr | 0.52 | 0.096 | 48.0%, 70.0% | 0, 0% |
| Lac/NAA | 0.48 | 0.058 | 45.8%, 60.0% | 0, 0% |
| **ADC** |  |  |  |  |
| Whole brain | 0.75 | 908.0 | 100%, 58.3% | 908.0, 58.3 |
| Cortex | 0.78 | 989.0 | 92.9%, 61.1% | 950.0, 55.6% |
| Hippocampus | 0.65 | 1051.4 | 64.3, 55.6% | 921.6, 27.8% |
| Thalamus | 0.57 | 927.6 | 50.0, 61.1% | 856.7, 27.8% |
| White matter | 0.76 | 821.9 | 92.9%, 61.1% | 802.5, 58.3% |
| Basal ganglia | 0.67 | 843.2 | 85.7%, 50.0% | 806.2, 33.3% |
| Cerebellum | 0.70 | 904.4 | 71.4%, 66.7% | 810.7, 38.9% |

### Classification of Evidence

The study provides Class IV evidence of MRS changes in anoxic brain injury after cardiac arrest.

### Standard Protocol Approvals, Registration, and Patient Consents

The Institutional Review Board approved this study, and consent was obtained for each patient by the legally authorized representative unless MRS was performed as part of clinical care.

### Data Availability Policy

Data will be shared at the request of other investigators for purposes of replicating procedures and results.

## Results

### Characteristics of Study Cohort

A total of 51 comatose patients after cardiac arrest underwent TTM, cEEG, MRI, and MRS. One patient was excluded due to presentation with a large posterior circulation stroke immediately before cardiac arrest, resulting in a total of 50 patients who underwent analysis. Of these, 14 had coma recovery, of whom 7 returned cognitively to baseline; all patients had benign EEGs (Table 1). Three patients who did not have coma recovery survived, remaining comatose at hospital discharge (CPC=4). One patient was awake, spontaneously moving extremities, and was noted to follow 1-step commands approximately 75% of the time (CPC=3). There were no meaningful differences in age, sex, or race. Patients who recovered were statistically significantly more likely to have intact corneal reflexes (100% vs 55.6%, p=0.002) and motor movements better than flexion (100% vs 11.1%, p<0.001).

MRS peaks were evaluated for effect of time elapsed to scan after cardiac arrest. Time to scan revealed a statistically significant positive correlation with NAA/Cr (r_s_=0.43 95% confidence interval = [0.17, 0.63], p=0.002) and a negative correlation with log(Lac/Cr) (r_s_=−0.40 [−0.11, −0.60], p=0.007).

### Persistent coma is associated with reduced NAA/Cr and increased Lac/Cr compared to patients with coma recovery and control subjects

PCG NAA/Cr was statistically significantly lower in patients who did not have coma recovery as compared to those who did (0.82 vs 1.26, p<0.001), with an AUC of 0.96 (Tables 1 and 2, Figure 2). The Optimal cut-off, defined as the value-maximizing Youden’s J statistic^17^, was 1.11 for a sensitivity of 85.7 and specificity of 86.5%. None of the patients with an NAA/Cr ratio under 0.98 recovered; 33% of patients between 0.98 and 1.2 recovered, and all patients above 1.2 recovered. PCG NAA/Cr was significantly lower in patients who died, as compared to those who survived (mean NAA/Cr ratio of 0.79 vs 1.20, p<0.001).

**Figure 2:**
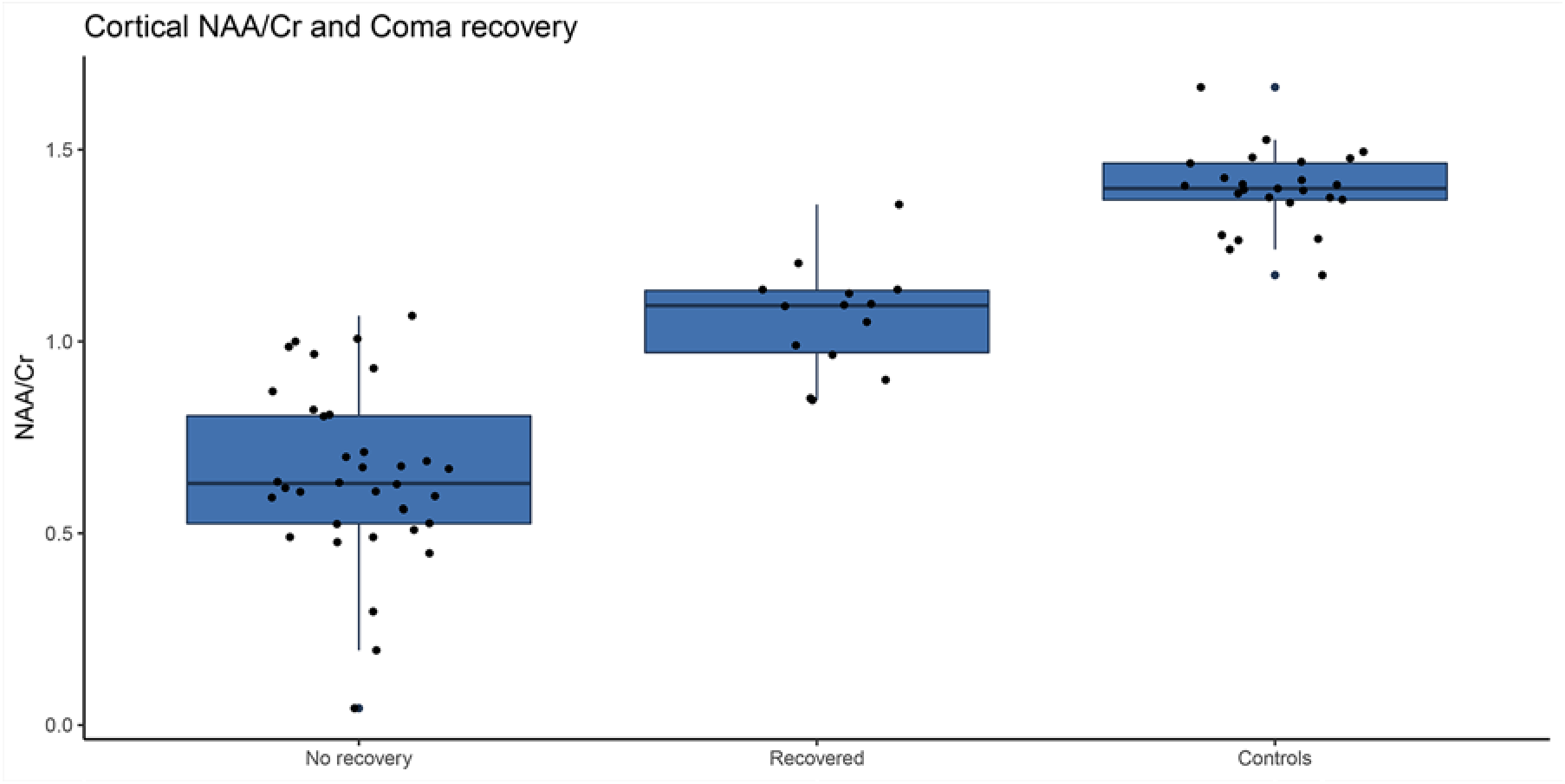
Posterior cingulate gyrus (PCG) NAA/Cr ratios in patient populations Boxplots of PCG NAA/Cr levels patients who had no coma recovery (did not follow commands), recovery (followed commands), and normative control subjects

PCG Lac/Cr was statistically significantly higher in patients who did not have coma recovery (0.39 vs 0.04, p<0.001, Table 2), with an ROC AUC of 0.91. There was one outlier survivor whose Lac/Cr was higher than expected (0.136). Although included for purposes of completeness, there were technical difficulties with the scan; a water reference was not obtained, which may have affected scaling ratios.

PCG Lac/Cr was also compared to peak serum lactate during admission, and serum lactate level in closest proximity to the MRS scan, all but one of which were within 72 hours. There was no statistically significant relationship between PCG MRS Lac/Cr peak and peak serum lactate (r_s_ =0.09, 95% CI [−0.20, 0.36]) or with serum lactate in the proximity of the MRS (r_s_ =0.17 [−0.12, 0.42]).

In the PWM, BG, and BS, there were smaller differences in NAA/Cr and Lac/Cr between patients who did or did not achieve coma recovery. Only differences in Lac/Cr peaks reached statistical significance before multiple comparisons correction in the WM and BG.

### Patients with coma recovery have reduced NAA/Cr as compared to control subjects

Patients were compared to control subjects with no history of brain injury or trauma. In patients without coma recovery, the following statistically significant changes were observed:

– Decrease in NAA/Cr (PCG [0.82 vs 1.40, p<0.001], PWM [1.48 vs 1.87, p<0.001], adjusted for multiple comparisons)
– Decrease in GABA (PCG [0.19 vs 0.27, p<0.001], PWM [0.20 vs 0.23, p<0.001]),
– Decrease in glutamate (PCG [0.97 vs 1.24, p<0.001])
– Decrease in Glx (PCG [1.74 vs 2.10, p=0.010], PWM [1.18 vs 1.36, p<0.001])
– Decrease in MI/Cr (PCG [0.55 vs 0.75, p<0.001])
– Increase in Lac/Cr (PCG [0.39 vs 0.073, p<0.001] and PWM [0.43 vs 0.22, p<0.001]])
– Increase in glutamine/Cr (PCG [1.12 vs 0.48, p<0.001], PWM [0.47 vs 0.30, p<0.001]).

In patients with coma recovery, NAA/Cr was significantly lower (PCG [1.26 vs 1.40, p<0.001], PWM [1.71 vs 1.87, p=0.008], in addition to a decrease in glutamate (PCG [1.12 vs 1.24, p=0.030]), and an increase in glutamine (PWM [041 vs 0.30, p=0.003]). No other peaks were statistically significantly different between the two populations, specifically, no difference in Lac/Cr was seen. These changes were still present in patients with coma recovery who had a cognitive return to baseline; NAA/Cr were lower in PCG (1.26 vs 1.40, p=0.001) and PWM (1.76 vs 1.87, p=0.035) and glutamine/Cr elevated (0.43 vs 0.30, p=0.020). These findings suggest that patients with coma recovery still sustained an anoxic injury, but not at the same scale as the non-recovered patients.

### Reduced MRI ADC signal intensity is associated with NAA/Cr reduction and lactate elevation

There were significant reductions in MRI ADC measures in patients who did not have coma recovery in the whole brain, cortex, hippocampus, and white matter, with the cortex being most significantly affected (Table 1). Correlation between PCG NAA/Cr and ADC values were examined for mean total brain ADC as well as mean ADC values for individual brain regions. There were significant correlations with the cerebellum, cortex, hippocampus, globus pallidus, putamen, BG (combined), and white matter regions. Correlation between NAA/Cr and ADC was linear (Figure 3), and it was strongest with cortical ADC values (r_s_=0.65 [0.45, 0.79], p<0.001). In comparison, there was a negative log-linear relationship between PCG Lac/Cr and cortical ADC (r_s_=−0.74 [−0.58, −0.84], p<0.001).

**Figure 3:**
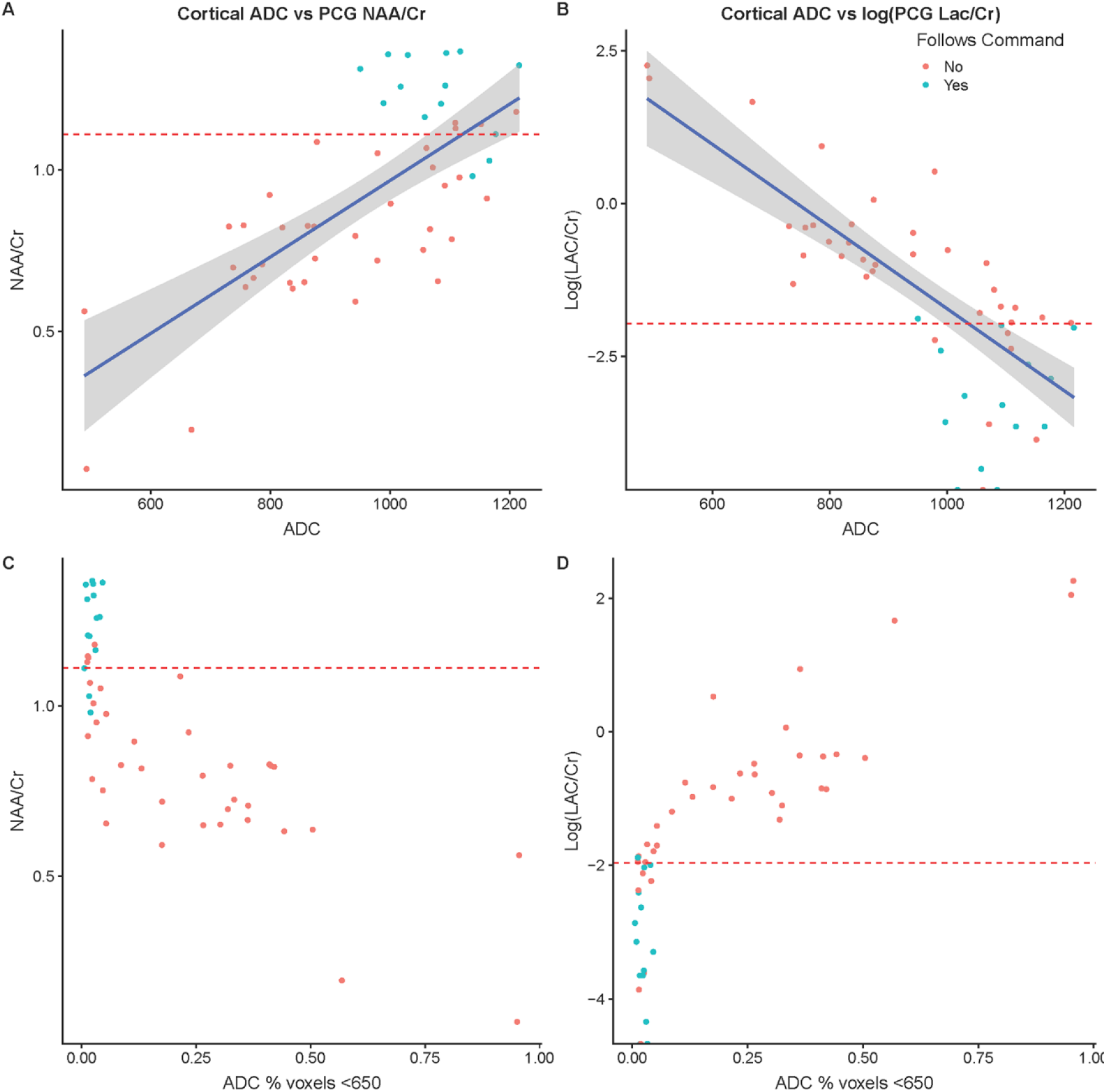
Relationship between cortical ADC vs posterior cingulate gyrus (PCG) NAA/Cr, PCG lactate, and coma recovery A) Linear relationship between cortical ADC and PCG NAA/Cr. The red line represents the optimal cut-off for recovery to follow commands. B) Inverse log-linear relationship between cortical ADC and PCG lactate. The red line represents the optimal cut-off for recovery to follow commands. The blue line represents cut-off with the specificity of recovery set to 100%. C) and D) Relationship between % of cortical voxels with ADC < 650 mm^2^/s and NAA/Cr (C) and log of PCG lactate (D).

**Figure 4:**
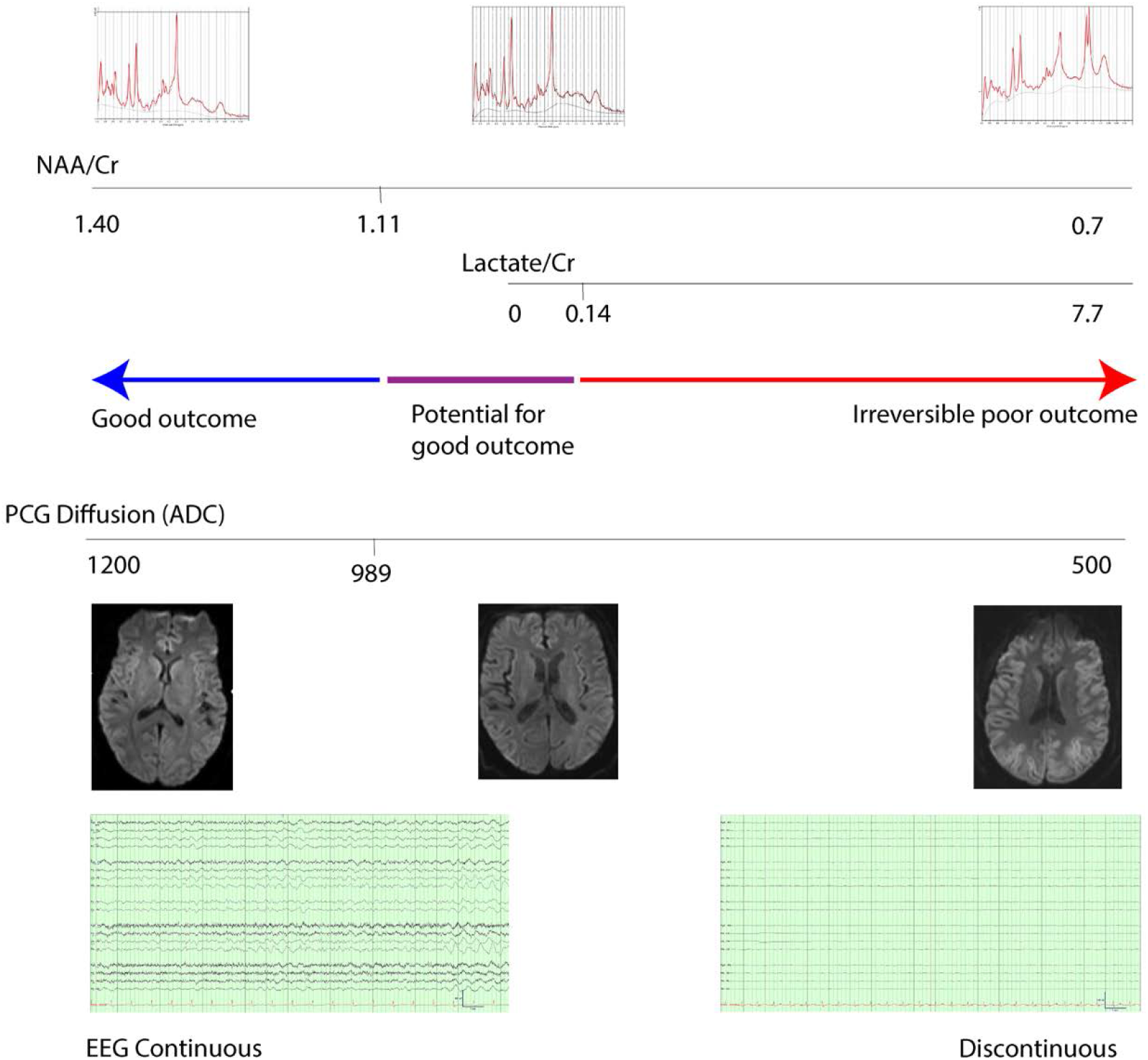
Illustration of the relationship between MRS, MRI, EEG, and outcome after cardiac arrest Summary of MRS and ADC peaks after TTM. NAA/Cr/Cr values higher than 0.98 and ADC values over 1071 resulted in universal good come with coma recovery. The appearance of lactate resulted in a poor outcome. Patients between these values (in purple) have the potential for coma recovery. For illustrative purposes, diffusion-weighted images are shown rather than ADC maps.

White matter ADC values were correlated to PWM Lac/Cr (r_s_=−0.46 [−0.35, −0.74], p=0.0044) with a trend towards correlation with PWM NAA/Cr (r_s_=0.28 [−0.0001, 0.52], p=0.052). Basal ganglia ADC values were correlated to NAA/Cr (r_s_=0.39 [0.008, 0.68], p=0.047) with a tend towards correlation with Lac/Cr (r_s_=−0.41 [−0.023, 0.69] but p=0.06 after multiple comparisons). Correlations between PWM and BG metabolites and ADC values from corresponding brain regions were otherwise not statistically significant.

### Continuous background on EEG is associated with higher NAA/Cr; suppressed or burst suppressed EEGs are associated with Lac/Cr production

Patients who had a continuous EEG background had higher NAA/Cr than patients with a discontinuous background (0.79 vs 1.18, p<0.001), and less likely to have a Lac/Cr over 0.14 (5 of 21 vs 26 of 29, p<0.001). Patients who did not recover from coma nearly always had discontinuous, burst suppressed, or completely suppressed EEGs whereas all patients who recovered had a continuous background on EEGs (Table 1). Reactivity to stimulation, though not tested or determinable in all patients, was more often present in patients who recovered (p<0.001). There were no differences in the number of patients who had status epilepticus, generalized periodic discharges, or myoclonus. All patients with suppressed EEG backgrounds at the time of MRS/MRI scans had elevated PCG Lac/Cr peaks of at least 0.331. All patients with burst suppressed EEGs had PCG Lac/Cr peaks of at least 0.167.

In patients who did not achieve coma recovery, Lac/Cr was lower in patients who experienced a seizure during any time of their hospitalization than patients who did not (0.452 vs 0.163, p=0.0048). No changes in lactate in relation to seizures were seen in patients who achieved coma recovery.

## Discussion

This study demonstrates the spectrum of metabolic changes on MRS in patients after anoxic brain injury; relationships with clinical, MRI, and electrographic markers of anoxic brain injury were as follows:

1. Poor coma recovery was associated with a decrease in PCG NAA/Cr by approximately 35% and elevated lactate production (in our cohort, Lac/Cr >0.14).
2. MRS assessment had the highest association with coma recovery in the PCG as compared to PWM, BG, or BS.
3. There is a linear relationship between cortical ADC values on MRI and PCG NAA/Cr and there is an exponential increase in PCG lactate with decreasing cortical ADC, (e.g. increasing cortical diffusion restriction).
4. Continuous background on EEG was associated with higher PCG NAA/Cr; EEGs that are suppressed or burst suppressed at the time of the MRS were universally associated with lactate production.

We observed a decrease in NAA/Cr values in patients with coma recovery as compared to normative control subjects. This decrease persisted even in patients who had cognitive return to baseline at bedside exam, though full neuropsychological evaluations were not performed. We hypothesize that this is likely the result of mild anoxic injury, rather than TTM itself exerting non-specific changes in the MRS spectra once the patient has been rewarmed. It is unclear whether these patients experienced more subtle longer-term cognitive or other deficits, despite their good functional outcome at hospital discharge. Previous studies have demonstrated long-term cognitive deficits in >40% of survivors of cardiac arrest.^18^ Interventions in addition to TTM to preserve structural and metabolic integrity after resuscitation to optimize neurological recovery, even in patients who appear to have excellent in-hospital outcomes, should be explored in future studies. Recent preclinical and clinical studies have suggested promising effects of noble gases^19^ and citicoline^20^ as neuroprotective agents.

Our cohort demonstrates a tight coupling between NAA/Cr and MRI ADC values. ADC measures the impedance of water molecule diffusion and assesses cell membrane integrity.^21^ NAA is a molecule found in the brain at high concentrations that is sensitive in a nonspecific manner to several neurologic disorders and has been postulated to be a marker of neuronal health and bioenergetic dysfunction.^22, 23^ The high degree of linear correlation of these two disparate measurements in the cortex further suggests that the cortex has the greatest sensitivity to anoxic injury, as compared to other sampled brain regions.

This study also highlights the importance of the appearance of a lactate peak. Although there was an overlap of NAA/Cr and ADC values in patients who had coma recovery vs those who did not, the appearance of a lactate peak in the PCG with a concentration of 0.14, on the other hand, was more consistently associated with the absence of coma recovery. Electrographically, complete background suppression or burst suppression, which are patterns that have been demonstrated to be associated with poor outcome in multiple studies^9, 24^, were consistently associated with a lactate peak. As such, the emergence of a lactate peak may represent a point of irreversible brain injury from which coma recovery is not possible.

As such, we hypothesize that MRS changes provide an in-vivo view of the major pathophysiological processes in anoxic brain injury that may occur at different severities (Figure 5). NAA/Cr may be representative of early injury that may be most sensitive but potentially nonspecific for poor outcome. Most patients with a mild decrease in NAA/Cr and ADC achieve coma recovery. Increasing severity of injury results in only a portion of patients recovering. Injury severe enough to cause an increase in lactate represents severe, potentially irreversible anoxic injury. Lactate levels as measured by MRS have been demonstrated to be a reliable marker of outcome after stroke.^25^ In a rat model, hypothermia induced neuroprotection-related metabolic changes were seen, including an increase in Lac and MI, and a decrease in Glu.^26^

The mechanism of elevated lactate peak in severe anoxic injury may be viewed in terms of the integrity of oxidative metabolism. Rapid depletion of ATP after cessation of cerebral perfusion causes failure in the membrane ATP-dependent Na/K pumps, resulting in a massive influx of Na, efflux of K, and membrane depolarization. This results in opening voltage-gated Ca channels resulting in large increases in intracellular Ca, activation of Ca-dependent K channels, resulting in further loss of selective membrane permeability.^27^ Lactate levels rise almost immediately occurs as a result of a switch to anaerobic glycolysis, which quickly returns to baseline if perfusion is restored and normal mitochondrial function is resumed. Accumulation of lactate after re-establishment of perfusion is due to secondary energy failure in which neurons that survive the initial insult develop energy depletion from mitochondrial failure^28, 29^; elevated MRS lactate at 48 hours is associated with cell death and microglial activation in animal models^30^, and the accumulation of lactate portended an extremely poor outcome in pediatric hypoxic-ischemic injury.^31, 32^ Failure to re-establish membrane gradient results in cytotoxic edema, cessation of cerebral perfusion, and neuronal transmission.

The failure of oxidative metabolism, as evidenced by lactate generation and its relationship to synaptic activity, may account for the consistent findings of either burst suppression or severe background voltage suppression seen in this cohort. Approximately 75-80% of the brain’s energy requirements are for signal processing, predominantly for the generation of action potentials and postsynaptic actions of neurotransmitters, predominantly glutamate.^33, 34^ The generation of EEG signals is due to a summation of local field potentials, the most important source of which is synaptic activity^35^. Varying levels of damage to the excitatory synapses have been demonstrated to recapitulate the various EEG abnormalities seen in postanoxic EEGs, including burst suppression and discontinuous low voltage recordings.^36^ Although it is likely that presynaptic ischemic failure is the initial cascade in inhibiting synaptic transmission in mild-moderate ischemia,^34^ severe and widespread failure in synaptic transmission is likely coupled to the impairment of oxidative metabolism.

One unexpected finding is the increase in glutamine in patients with poor outcomes. Glutamate and glutamine are preferentially used as metabolic and cataplerotic substrates in the Krebs cycle during anoxia and ischemia. They may also function as reservoirs to protect against post-ischemic reduction in cardiac output by maintaining metabolic intermediates. The biosynthesis of glutamine from glutamate amidation is catalyzed by the enzyme glutamine synthetase (GS); after the release and reuptake of glutamate into neurons and glia, glutamate is catalyzed back to glutamine and ammonium by mitochondrial phosphate-activated glutaminase (PAG). GS activity increases in response to acute hypoxic-ischemic nervous system injury in children and other compensatory mechanisms prevail in the case of chronic hypoxic-ischemic insults.^37^ In cell culture models of hypoxia, PAG activity is inhibited, potentially by the acidic pH induced by lactic acidosis.^38, 39^ The combination of increased glutamine synthetase activity and inhibition of phosphate-activated glutaminase may play a rescue role in preventing glutamate-induced injury. Separating the glutamine and glutamate resonances remains challenging, and further studies are required to confirm these findings.

Our study is in agreement with a large European study that demonstrated that NAA/Cr ratios measured in the pons and thalami were significantly lower in patients with unfavorable as compared to favorable outcomes for MRI/MRS obtained between 7 and 28 days after cardiac arrest.^3^ Differences from our study include that their study reported only MRS values of the thalamus and pons, included patients both patients who did and did not undergo TTM, and included only patients who had survived for 7 days after cardiac arrest. PCG is more sensitive for identifying MRS changes when compared to the PWM, BG, and BS. Furthermore, we demonstrate a robust correlation between single voxel spectra of the PCG and both cortical and whole-brain ADC, suggesting that spatial sampling bias may be less relevant as loss of perfusion as well as reperfusion is global. Future studies with multivoxel spectroscopy will help to fully elucidate regional variations.

### Study Limitations

The sample size is too small to develop a multimodal outcome prediction model. This also likely explains the lack of association between coma recovery and status epilepticus, generalized periodic discharges, or myoclonus. The cut-off values are thus representative of only our dataset. Voxel selection did not target regions of greatest ADC or T2 changes on MRI scans, potentially decreasing MRS effect size. However, this minimized variability in signal/noise characteristics of different brain regions and allowed the generalizability of findings across subjects. As MRS scans were only obtained with a clinically necessary MRI scan, there is variability in the timing of the image acquisition and potential selection bias. Although there were correlations between time elapsed to scan and MRS peaks, this may reflect a strong bias for patients with clinical suspicion of poor outcome to be scanned earlier in their course of hospitalization. Previous MRS studies in stroke^25^ and neonatal hypoxic-ischemic injury^40^ demonstrated a sustained decrease in NAA and a more transient increase in Lac. Without longitudinal MRS data, we are unable to determine the nature of MRS peak changes over time. Cortical ADC values of survivors are higher than typically seen in other studies, likely due to our selection of a more conservative cut-off value of 2000 mm^2^/s to remove artifacts^41^, likely resulting in the inclusion of high ADC CSF voxels. Nonetheless, our results remained robust across a range of other cut-off values (1000 mm^2^/s and 1500 mm^2^/s).

NSE and SSEPs were not routinely obtained on every patient, and a pupillometer was not utilized. We assessed cognitive return to baseline to the patient’s estimated premorbid baseline on bedside exam. A detailed neuropsychological evaluation, which was not performed, would be required to fully assess residual deficits. WLST was performed on a case-by-case basis and may potentially conflict with objective prognostic measures and treating clinicians were aware of the MRS results. The choice of coma recovery defined as following commands, rather than CPC, as the primary clinical outcome, minimized the effect of these variabilities.

As with any testing modality, care must be taken in avoiding overreliance on a single measure including the MRS, as demonstrated by one patient who underwent 2 MRS scans. A female in her 40’s sustained a PEA arrest, thereafter experienced myoclonic status epilepticus. MRI/MRS were obtained 4 days and 7 days after cardiac arrest; while neither scan revealed lactate or ADC changes, NAA/Cr of the basal ganglia decreased 20.9% between the two scans. The patient eventually made a full recovery, without chronic seizures or observable cognitive deficits on long-term follow-up.

## Conclusions

This study shows marked differences in several MRS detectable metabolites in cardiac arrest patients undergoing TTM who achieved coma recovery, as compared to those who did not. NAA/Cr and diffusion restriction changes are linearly correlated. The increasing severity of the injury is associated with the emergence of a rapidly increasing lactate peak, which was associated with either burst or complete suppression on EEG, and from which no patients recovered. Metabolic changes may be seen after TTM as compared to normative controls even in patients with presumed excellent recovery. This suggests that some amount of neuronal injury and breakdown in cellular microarchitecture may be tolerated, whereas compromise to aerobic oxidative metabolism represents severe, potentially irreversible anoxic brain injury.

## Appendix 1: Authors

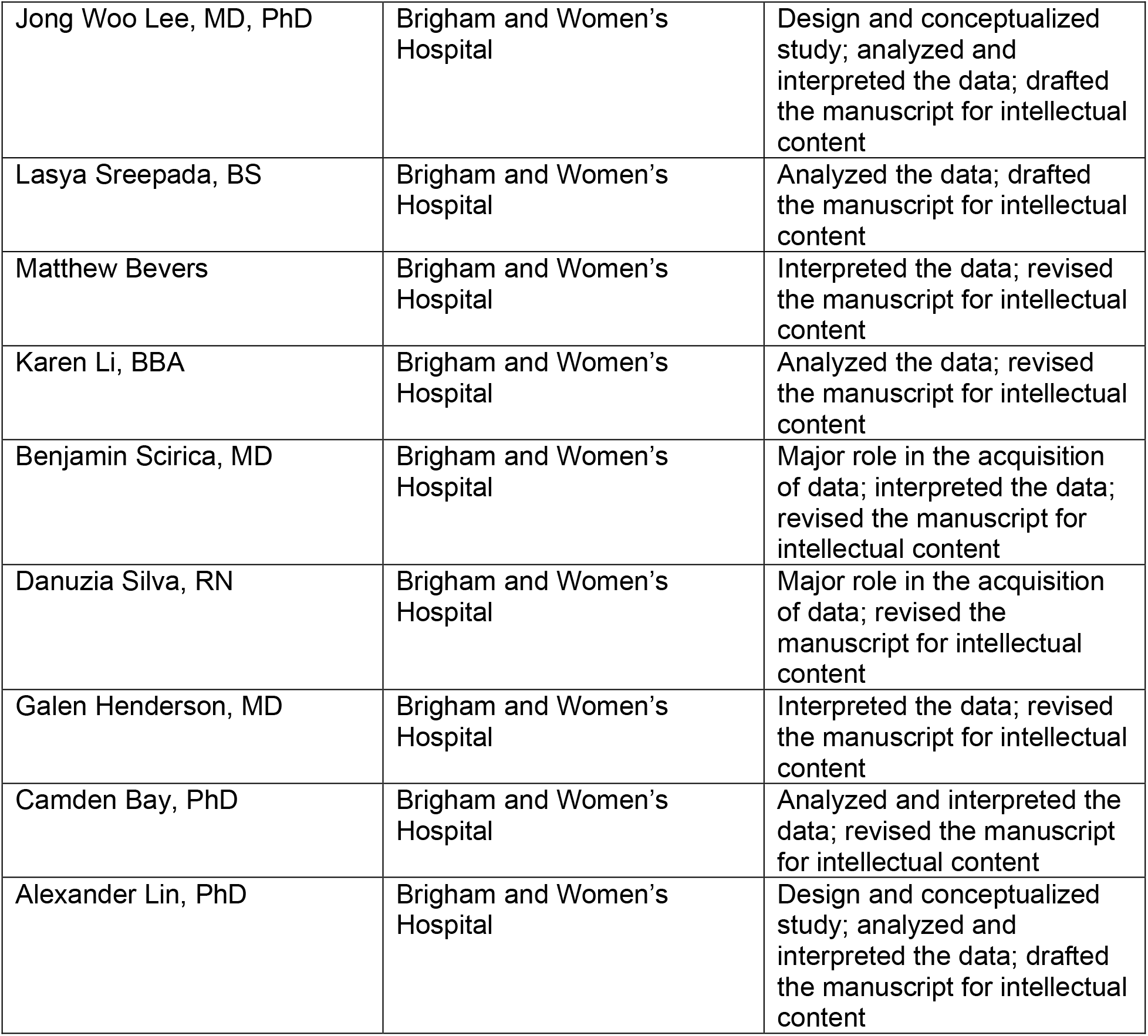

## Notes

**Study Funding**: The authors report no targeted funding

**Disclosures**: Jong Woo Lee has performed contract work for Teladoc and Bioserenity, he was the site PI for Engage Therapeutics, he has received research funding from the NINDS, he is the co-founder of Soterya, Inc; Lasya Sreepada reports no disclosures relevant to the manuscript; Matthew Bevers is supported by grants from the American Academy of Neurology and National Institute of Neurologic Disorders and Stroke, he reports research funding and personal fees from Biogen, outside the scope of the current work; Karen Li reports no disclosures relevant to the manuscript; Benjamin Scirica reports Institutional research grant to Brigham and Women’s Hospital from AstraZeneca, Eisai, Novartis, and Merck, consulting fees from AbbVie, Allergan, AstraZeneca, Boehringer Ingelheim, Covance, Eisai, Elsevier Practice Update Cardiology, GlaxoSmithKline, Lexicon, Medtronic, Merck, NovoNordisk, Sanofi, and equity in Health [at] Scale, is a member of the TIMI Study Group which has received institutional research grant support through Brigham and Women’s Hospital from: Abbott, Amgen, Aralez, AstraZeneca, Bayer HealthCare Pharmaceuticals, Inc., BRAHMS, Daiichi-Sankyo, Eisai, GlaxoSmithKline, Intarcia, Janssen, MedImmune, Merck, Novartis, Pfizer, Poxel, Quark Pharmaceuticals, Roche, Takeda, The Medicines Company, Zora Biosciences; Danuzia Silva reports no disclosures relevant to the manuscript; Galen V. Henderson reports no disclosures relevant to the manuscript; Camden Bay reports no disclosures relevant to the manuscript; Alexander Lin is a consultant for Agios Pharmaceuticals, Biomarin Pharmaceuticals, Moncton MRI and is cofounder of BrainSpec. He receives research funding from NINDS, NIA, Department of Defense, and the Alzheimers Association.

### Competing Interest Statement

The authors have declared no competing interest.

### Author Declarations

The Institutional Review Board of Mass General Brigham which oversees all human-subject research conducted by a Mass General Brigham-affiliated investigator approved this study, and consent was obtained for each patient by the legally authorized representative unless MRS was performed as part of clinical care.

